# Who is at the highest risk from COVID-19 in India? Analysis of health, healthcare access, and socioeconomic indicators at the district level

**DOI:** 10.1101/2020.04.25.20079749

**Authors:** Arindam Nandi, Ruchita Balasubramanian, Ramanan Laxminarayan

**Author notes:** **Corresponding author contact information:** 962 Wayne Avenue, Suite 530, Silver Spring, MD 20910-4433.

## Abstract

**Introduction:** Despite measures such as travel restrictions and lockdowns, the novel coronavirus (SARS-COV-2) is projected to spread across India. Considering that a vaccine for COVID-19 is will not be available soon, it is important to identify populations with high risk from COVID-19 and take measures to prevent outbreaks and build healthcare infrastructure at the local level.

**Methods:** We used data from two large nationally representative household surveys, administrative sources, and published studies to estimate the risk of COVID-19 at the district level in India. We employed principal component analysis to create an index of the health risk of COVID-19 from demographic and comorbidity indicators such as the proportions of elderly population and rates of diabetes, hypertension, and respiratory illnesses. Another principal component index examined the socioeconomic and healthcare access risk from COVID-19, based on the standard of living, proportion of caste groups, and per capita access to public healthcare in each district.

**Results:** Districts in northern, southern and western Indian states such as Punjab, Tamil Nadu, Kerala, and Maharashtra were at the highest health risk from COVID-19. Many of these districts have been designated as COVID-19 hotspots by the Indian government because of emergent outbreaks. Districts in eastern and central states such as Uttar Pradesh, Bihar, and Madhya Pradesh have higher socioeconomic and healthcare access risk as compared with other areas.

**Conclusion:** Districts at high risk of COVID-19 should prioritize policy measures for preventing outbreaks, and improving critical care infrastructure and socioeconomic safety nets.

## 1. Introduction

The novel coronavirus (SARS-COV-2) in India is projected to spread widely across a population^1^. Although early measures were implemented including stopping flights to China in January 2020, quarantining suspected cases coming into the country, and imposing a total lockdown on March 24, 2020, there is nevertheless widespread transmission of SARS-COV-2 around the country. As of April 24, 2020, there were 2,736,979 total confirmed cases and 192,125 deaths reported from the coronavirus infectious disease (COVID-19)^2^. Because testing for COVID-19 remains low in India (a rate of.023 per 1000 population as of April 19, 2020), these numbers likely represent only a small proportion of the true underlying burden of disease^3^. Even accounting for this undercount, the 42 day lockdown is projected to push out the epidemic curve in India^4^. However, the removal of the lockdown on May 3, 2020 will increase transmission, the extent of which will be determined by post-lockdown measures.

An important question relates to which parts of India are likely to bear the greatest burden of illness. SARS-COV-2 is likely to be transmitted in different places at different times, but the risk of severe disease and mortality, which are determined by age, gender, risk factors including obesity, hypertension and diabetes, varies greatly across the country. The aim of this paper was to estimate the district-level risk of severe-COVID-19 and mortality based on a principal component analysis.

## 2. Methods

### 2.1. Data Sources

We used data from the National Family Health Survey of India 2015-2016 (NFHS-4)^5^ which was a cross-sectional survey of 601,509 households and 2.87 million individuals from all states and union territories. The survey collected data on various socioeconomic, demographic, health, and family planning indicators. For 699,686 women of age 15-49 years and 112,122 men of age 15-54 years covered in the survey, additional measures of anthropometry and biomarkers related to anemia, hypertension, and diabetes were collected‘^-^’. NFHS-4 is the most recent source of such biomarker data at the district level in India.

We obtained additional information on the number of public health facilities in each district - primary health subcenters, primary health centers, community health centers, sub-divisional hospitals, and district hospitals - from the National Rural Health Statistics of India, 2017 (NRHS 2017).^6^ From the National Sample Survey of India 2017-2018 (NSS 75^th^ round), we also obtained the state level proportions of hospitalized patients who chose public vs. private healthcare providers during the year preceding the survey.? We estimated the population size in each district by using population distribution data from NFHS-4 along with the 2020 national population estimate for India from the United Nations.^8^

### 2.2. Principal Component Analysis

We used principal component analysis (PCA) which is a widely used method for reducing dimensionality through orthogonal linear transformation of the underlying data.^9^ In a multivariate setting, the orthogonal unit eigenvectors calculated from the covariance matrix of the data are known as the principal components. We included the following district level proportions – estimated from NFHS-4 – in our PCA: (i) 70-79 year old population, (ii) population of age 80 years and above, (iii) adults with diabetes or risk of diabetes, (iv) and adults with high blood pressure. These indicators were chosen following a large study of 72,314 COVID-19 patients in Wuhan which identified high case fatality rates among the elderly and those with diabetes, hypertension, and other health conditions.^10^

Diabetes and diabetes risk in our data were defined as blood sugar level of higher than 140 milligrams per deciliter (glucose tolerance). High blood pressure was defined as systolic pressure of over 160 and/or diastolic pressure of over 100. Blood pressure and sugar data were available for 15-49 year old women and 15-54 year old men. While the prevalence rates of diabetes and hypertension in the elderly are likely to be higher than these younger-age estimates, we assumed that the relative rates across districts would remain similar. We considered the first principal component, which captures most of the variance in the data, as an index of health risk of the district population from COVID-19.

We estimated another composite index of the socioeconomic risk, along with access to public and private healthcare, at the district level. The following district proportions (from NFHS-4) were included in this PCA: (i) socioeconomically disadvantaged scheduled caste (SC) or scheduled tribe (ST) groups, (ii) rural population, and (iii) population belonging to the lowest wealth quintile. Wealth quintiles were created from a composite index of possession of assets such as TV, radio, bicycle, and a car, along with indicators of quality of housing construction, and availability of toilets, electricity, and clean drinking water at the household.^11,12^ Three other covariates were included from NRHS 2017: (iv) inverse of the population size served per primary health center, (v) inverse of the population size served per community health center, and (vi) inverse of the population size served per sub-divisional hospital.^6^ NFHS-4 data covered all 640 districts of India during 2015-2016, and those were later divided into 698 districts in NHRS 2017. We matched the NRHS 2017 districts backwards with NFHS-4 districts. Districts which were completely urban, such as Kolkata and Mumbai, were not covered by the NRHS, and therefore excluded from this analysis.

Finally, from NSS 75^th^ round survey data, we included (vii) the proportion of hospitalized patients who chose a private healthcare provider in each state. We considered the first principal component extracted from the model as a composite index of the socioeconomic and healthcare access risk related to COVID-19 at the district level.

### 2.3. Sensitivity analysis

We conducted sensitivity analysis for the health risk index in two ways. First, we included additional indicators – prevalence or incidence rates as available – of chronic obstructive pulmonary disease, asthma, cancer (all causes), and ischemic heart disease at the state level among the covariates of the PCA model (Table 1). These data were not available at the district level. Second, instead of PCA which estimates factor loads (weights) of the covariates based on the covariance matrix, we constructed the health risk index using a simple weighting scheme based on the Wuhan study.^10^ We standardized the four district level indicators from the original model and weighted them with the case fatality rate of COVID-19 from Wuhan. The health risk index was calculated as the weighted sum. Table 2 presents the weighting scheme.

**Table 1:** Principal Component Analysis comorbidity indicators at the state level.

| **Chronic disease type** | **Indicator (at the state level)** | **Range** | **Source** |
| --- | --- | --- | --- |
| Ischemic Heart Disease | Prevalence per 100,000 | 1,017-2,793 | <sup>19</sup> |
| Diabetes | Prevalence per 100 adults >20 years old | 5.3-13.1 | <sup>20</sup> |
| Asthma | Prevalence per 1000 | 30-5,783 | <sup>21</sup> |
| Chronic Obstructive Pulmonary Disease | Prevalence per 1000 | 23-9,393 | <sup>21</sup> |
| Cancer | Crude Annual Incidence Rate | 53.9-135.3 | <sup>22</sup> |
| Hypertension | Prevalence per 100 | 12.8-40 | <sup>23</sup> |

Note: Data on these comorbidity indicators were available at the state level from published studies.

**Table 2:**
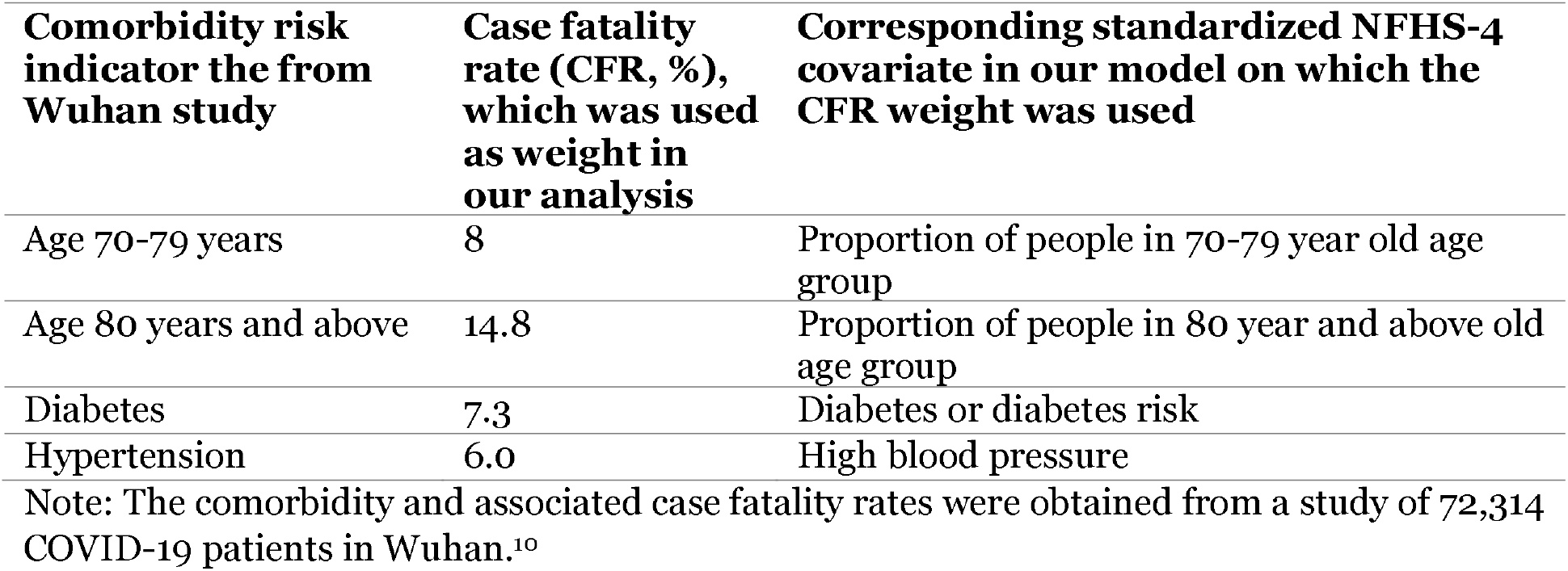
Weighting scheme of covariates based on Case Fatality Rate in Wuhan, China.

## 3. Results

Results from the health risk PCA are presented as a district heat map in Figure 1, while those from socioeconomic and healthcare access risk PCA are presented similarly in Figure 2. The underlying PCA factor loadings and coefficients are presented in Tables 3 and 4 respectively. We divided the estimated risk index into 15 equal categories - darker colors indicate higher risk to the district from COVID-19. On April 15, 2020, the Indian government designated 170 districts with a high number of confirmed COVID-19 cases as hotspot districts.^13^ These districts are indicated with blue colored dots on all maps.

**Figure 1:**
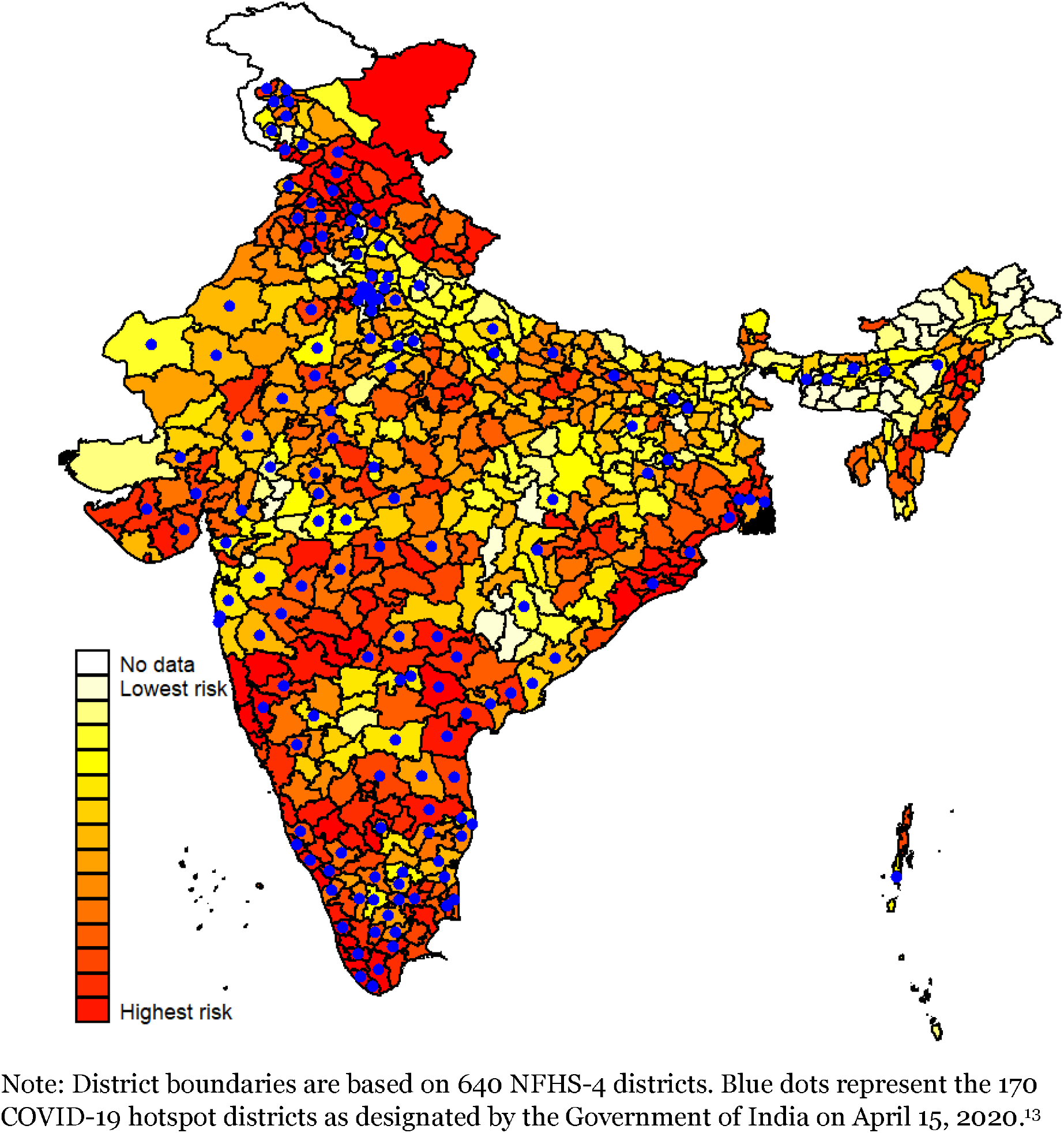
Composite (principal component) index of health risk from COVID-19 at the district level in India.

**Figure 2:**
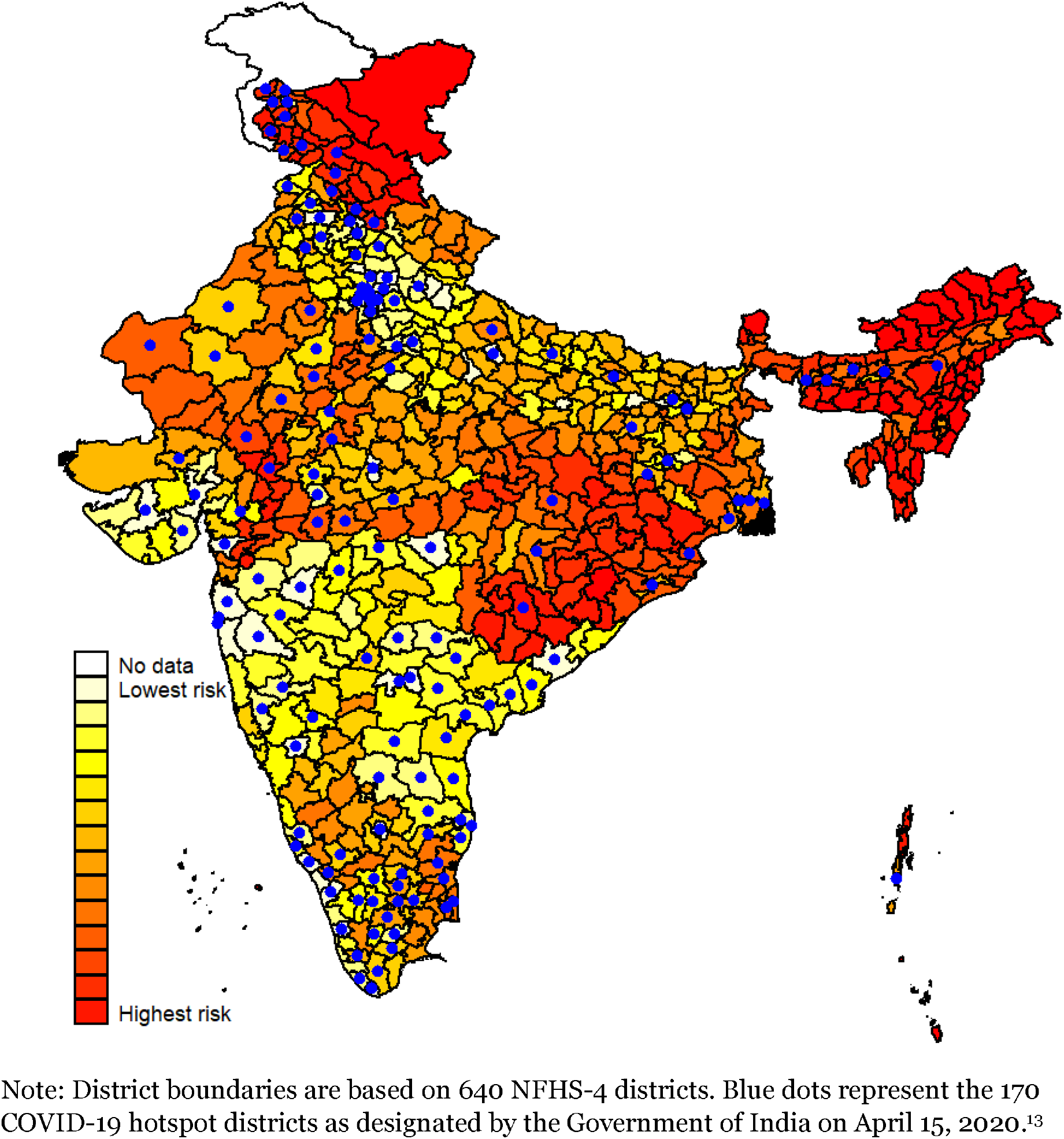
Composite (principal component) index of socioeconomic and healthcare access risk from COVID-19 at the district level in India.

**Table 3:** Results of principal component analysis for health risk index (first principal component)

| **Variable** | **Factor loadings (Factor 1)** | **Uniqueness** | **Scoring coefficient** |
| --- | --- | --- | --- |
| Proportion of 70-79 year old population | 0.93 | 0.14 | 0.51 |
| Proportion of 80 years and above old population | 0.91 | 0.17 | 0.50 |
| Proportion of population with diabetes or diabetes risk | 0.36 | 0.87 | 0.20 |
| Proportion of population with high blood pressure | 0.06 | 1.00 | 0.03 |

Note: Data were from NFHS-4 (2015-2016) survey. Regression based scoring coefficient was used.

**Table 4:** Results of principal component analysis for socioeconomic and healthcare access risk index (first principal component)

| **Variable** | **Factor loadings (Factor 1)** | **Uniqueness** | **Scoring coefficient** |
| --- | --- | --- | --- |
| Proportion of population in the lowest wealth quintile | 0.13 | 0.98 | 0.05 |
| Proportion of scheduled caste and scheduled tribe population | 0.73 | 0.47 | 0.30 |
| Proportion of population in rural areas | 0.36 | 0.87 | 0.15 |
| Inverse of Population size served by each primary health center | 0.84 | 0.29 | 0.35 |
| Inverse of Population size served by each community health center | 0.78 | 0.39 | 0.32 |
| Inverse of Population size served by each sub-divisional hospital | 0.03 | 1.00 | 0.01 |
| Proportion of hospitalized patients served by private health facilities | -0.65 | 0.57 | -0.27 |

Note: Data were from NFHS-4 (2015-2016) survey, National Rural Health Statistics 2017, and NSS 75<sup>th</sup> round survey (2017-2018). Regression based scoring coefficient was used.

## 4. Discussion

While several pharmaceutical companies are concurrently trying to speed up the development of a SARS-COV-2 vaccine, the timeline of such an intervention is set at more than 1 year. There is a need for identifying populations at high risk of COVID-19 outbreaks to preemptively implement solutions to mitigate this risk^14^. Previous studies have developed computational tools to assess the risk of COVID-19 outbreaks in countries worldwide outside of China, but these tools primarily assess risk based on a country’s connectivity to China, the efficacy of its entry screening, and the efficacy of its control measures^15^. These tools have since lost relevance in light of emerging travel restrictions and lockdowns which have shifted the epicenters of the outbreak.

Newer studies are identifying risk of SARS-COV-2 transmission at subnational levels, such as the counties of the United States, where they identified rural regions as being particularly susceptible to transmission^16^. A similar risk assessment has yet to be conducted in India which harbors several risk factors for COVID-19 including high burdens of diabetes and cancer^17,18^. We identified the Indian districts that are likely to experience COVID-19 outbreaks earlier than others.

Our results show that districts in northern, southern and western Indian states such as Punjab, Tamil Nadu, Kerala, and Maharashtra are at the highest health risk from COVID-19 (Figures 1,3, and 4). The pattern of higher health risk in these districts is especially pronounced in Figure 3 which presents the most detailed PCA including additional chronic disease indicators at the state level. The populations in these districts have higher proportions of the elderly and higher rates of chronic diseases such as diabetes and hypertension than the rest of India. In comparison, districts in eastern and central states including Uttar Pradesh, Bihar, and Madhya Pradesh have higher socioeconomic and healthcare access risk than other districts (Figure 2). The populations in these districts have lower standard of living, higher proportions of rural, scheduled caste, and scheduled tribe groups, and lower levels of access to public healthcare facilities as compared with the rest of India. While overall health risk of COVID-19 may be lower in these districts as compared with districts from northern, southern and western Indian states, those who contract the disease in these districts are likely to suffer substantially due to poverty and lack of access to care.

**Figure 3:**
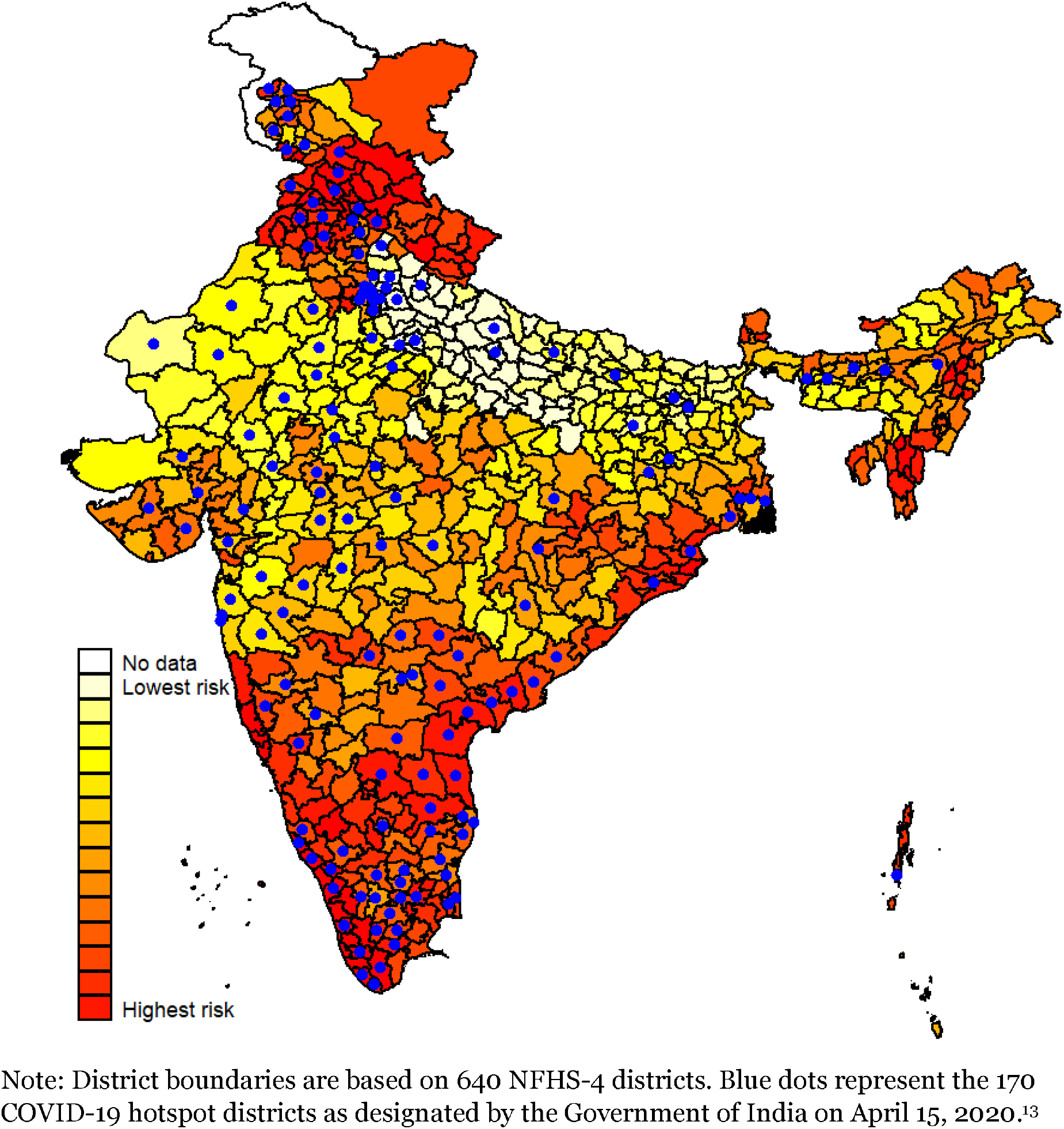
Sensitivity analysis – composite (principal component) index of health risk from COVID-19 at the district level in India, including additional state level indicators of chronic disease rates.

**Figure 4:**
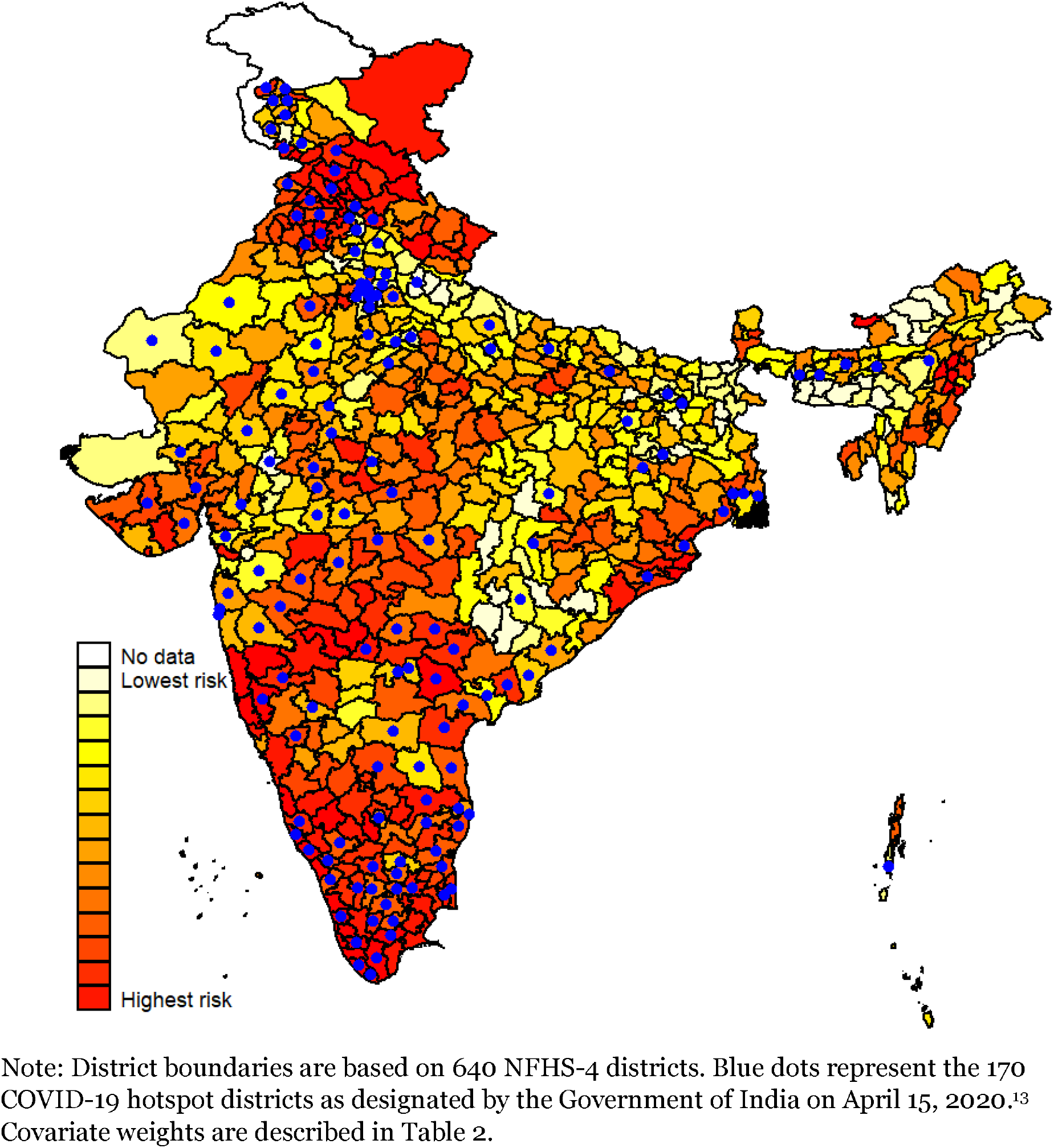
Sensitivity analysis – index of health risk from COVID-19 at the district level in India, with covariate weights based on case fatality rates from the Wuhan study.

On April 15, 2020, the Union Health Ministry of India identified 170 districts across 25 states as COVID-19 hotspot districts.^13^ These districts contributed to either more than 80 percent of the total COVID-19 cases in India or more than 80 percent of the total COVID-19 cases in each state. Districts that reported high growth rates of infection were also considered as hotspots. A substantially large number of hotspot districts matched well with the districts which we identified to be at high health risk (Figure 1,3,4).

Our findings can be used by policymakers to guide appropriate response, but must be interpreted carefully, as there may be additional factors such as population density and local policy measures which may affect the risk but could not be included in this analysis. Nevertheless, these findings provide a forecast of the potential hotspots of transmission that can inform preemptive responses to alleviate the repercussions of a sustained COVID-19 outbreak.

## Data Availability

Data are publicly available from Indian government sources and published studies.

https://data.gov.in/

## Conflict of interest

None declared

## Notes

### Competing Interest Statement

The authors have declared no competing interest.

### Funding Statement

No external funding was received for this work.

### Author Declarations

This study used publicly available data from secondary sources. No separate ethics clearance was necessary.

